# Development and validation of the Behavioral Health Acuity Risk model: a predictive model for suicide prevention through clinical interventions

**DOI:** 10.1101/2022.12.21.22283796

**Authors:** Varun Digumarthi, Heather E Strange, Heather B Norman, Derek Ayers, Raj Patel, Karen E Hegarty

## Abstract

Common suicidal ideation screening tools used in healthcare settings rely on the willingness of the patient to express having suicidal thoughts. We present an automatic and data-driven risk model that examines information available in the medical record captured during the normal course of care. This model uses random forests to assess the likelihood of suicidal behavior in patients aged seven or older presenting at any healthcare setting. The Behavioral Health Acuity Risk (BHAR) model achieves an area under the receiver operating curve (AUC) of 0.84 and may be used on its own or as a component of a comprehensive suicidal behavior risk assessment.

## Introduction

Suicide was among the top four leading causes of death in the United States in 2020 for those between the ages of ten and forty-four years old, and the twelfth leading cause of death among all ages (1). The economic impact is estimated at more than $4.75 billion per year in the United States (2).

Perhaps surprisingly, many decedents of suicide receive some suicide risk assessment or contact with their healthcare provider prior to suicide (3,4). Unfortunately, current standards of care and most existing instruments for suicide risk assessment have been unable to accurately predict risk with high accuracy (5,6,7), and many patients who are assessed for suicidal ideation deny suicidal intent even within one week of completion of suicide (3). For example, the Columbia-Suicide Severity Rating Scale (C-SSRS) (8) was introduced in 2011 and has come to be widely considered as the gold standard for suicide risk assessment. As with other tools relying on self-reported information, its utility is limited by the respondent’s willingness to provide truthful answers about intent to commit suicide or self-harm. It has been shown to identify patients at risk at a rate only moderately higher than random, having an area under the receiver operating curve (AUROC) of 0.65 (6), and that the assessment itself is susceptible to navigation errors and misinterpretations of question intent (7).

Various computer-aided models have been developed to identify high risk individuals. Tiet et al. developed a decision tree model for individuals with substance use disorder and recent suicidal ideation which evaluates prior suicide attempts, substance use, suicidal ideation, and violent behavior of patients to discriminate between those who will have a future suicide attempt and those who will not (9). Kessler et al. explored regression models for predicting suicide within a population of male non-deployed Regular U.S. Army soldiers having outpatient mental health specialty visits (10). One of the challenges faced by previous researchers is the low prevalence rate of suicide within a general population, which poses difficulty for many statistical and machine learning modeling techniques which suffer under the constraints of a highly imbalanced data set. Typically, the sensitivity and specificity of preceding models has been limited (11,12), which is a barrier to widespread adoption in clinical settings.

The objective of the work described here was to develop and validate a predictive model, which we refer to as the Behavioral Health Acuity Risk (BHAR) model, utilizing machine learning to assess likelihood of an individual to complete suicide. The model is designed for any individual aged 7 and older presenting at any healthcare setting for any cause regardless of previous history of suicidal ideation or attempt. The BHAR model generalizes to a wide population and enables behavioral health care team members to direct resources in the most meaningful way to prevent potential harm. It was developed to be used either on its own or in conjunction with a screening instrument such as the aforementioned C-SSRS to provide a comprehensive assessment of risk through both self-reported information and automated machine learning driven assessment. We compare the performance of the BHAR model to that of the C-SSRS screening tool and evaluate the effectiveness of the BHAR model in identifying patients at risk over the C-SSRS which is reliant on self-reported information.

## Materials and methods

### Data collection

All research was conducted according to a protocol approved by the Novant Health Institutional Review Board and in accordance with the ethical standards of the Helsinki Declaration of 1975. Data was sourced from existing historical electronic health records housed in the health system’s Epic Clarity database (13) and included any patient having been seen by a Novant Health provider between January 1, 2016 and August 1, 2020. Novant Health is an integrated health system located in the southeastern United States consisting of 15 medical centers and a medical group of >1,800 physicians at >800 ambulatory clinics (14). In addition to these health record data, mortality records from the state of North Carolina Department of Health and Human Services (15) during the same period were used to identify suicide completions which the health system may not otherwise have documented in the medical record. The data set included all encounters for all patients aged 7 years or older. In total, data from 2,884,214 patients were used for development of the model, with the model trained and tested at the encounter level. On average there were 15 encounters per patient.

### Feature and label construction

A set of 81 variables were investigated for predictive value in identifying patients who would later have completed suicide with a prediction horizon of 1 week from a given encounter. The predictors included patient demographic and socio-economic characteristics such as age at encounter, gender, race, ethnicity, and marital status. Behavioral characteristics and social determinants of health included access to firearms, feelings of hopelessness or helplessness, use of tobacco, use of alcohol, use of illegal drugs, volume of alcohol consumed per week, receiving routine care through a primary care physician, and prior history of self-injury. Total scores extracted from behavioral assessments such as the Patient Health Questionnaire-9 (PHQ-9) (16) and Generalized Anxiety Disorder-7 (GAD-7) (17) were also included. Predictors related to patient utilization history included prior outpatient visit to a behavioral health specialty clinic in the last 3 days, 30 days and 365 days, as well as having an inpatient admission into behavioral health units in the last 3 days, 30 days and 365 days. Presence or absence of diagnoses such as anxiety disorders, post-traumatic stress disorder (PTSD), and substance use disorders were included as binary variables. For diagnosis related predictors, two features were created for each diagnosis; first, presence or absence of a history of a diagnosis in the patient’s lifetime, and second, if the patient was diagnosed for the first time in the last 6 months. In addition to these discrete diagnosis related predictors, the calculated Charlson Comorbidity Index (CCI) score was also included as a predictor. Predictors related to prescribed medications include, for example, antipsychotics, antidepressants, benzodiazepines, and hypnotics. Medication features utilized a six-month lookback of medications reconciled or prescribed during both inpatient and outpatient clinical settings and were indicated as binary variables.

### Model training and evaluation

A random forest model (18) was selected to model the likelihood of suicide due to suitability to this problem and alignment to technical deployment constraints within the EHR platform. A random forest consists of many independently built decision trees. Each tree is built from a randomly selected subset of features and by bootstrap aggregation of training data. This helps to prevent overfitting without compromising on accuracy. Moreover, random forest models can be natively hosted in the EPIC EHR platform (13), enabling the risk score to calculate in near real time and be immediately visible to clinical staff. As mentioned previously, due to the low prevalence of suicide within the general population this dataset is highly imbalanced. Balancing can be achieved by various techniques, and we chose to employ the down sampling method. Down sampling the majority class to achieve a 1:16 ratio was found to be optimal through a hyperparameter search. That is, all the positive cases were retained, and then negative cases were randomly sampled into the data set until the desired ratio was met. The resulting data set was then split into training and testing encounters with an 80% train to 20% test split.

Hyperparameter tuning was performed using a grid search method with 3-fold cross validation having max tree depth between 15 and 30, max bins between 16 and 32, number of trees between 20 and 200 and feature subset strategy between 10 and 50. For binary classification tasks, model discrimination ability is frequently evaluated using techniques such as the Area Under the Receiver Operating Curve (AUROC) or the Area Under the Precision-Recall Curve (AUPRC). As even the down sampled data set was imbalanced in terms of outcome label, and because a prioritization is placed on the identification of positive cases, we chose to optimize the model using the AUPRC. Packages from pyspark machine learning were used to create a pipeline for training and testing. Matplotlib was used for creating visualizations. All extraction, analyses, transformations, and training were performed using spark architecture on 8 Azure-hosted compute-optimized standard F4s with 4-core CPUs and 8 GB RAM. For model explainability and interpretability, we examine feature importance which captures the effect that each variable imparts to calculating risk. Feature importance is calculated using Gini impurity.

## Results and Discussion

The final model incorporates 69 features. Table 1 details these features and their corresponding importance in calculating the risk score.

**Table 1.** Model features, description, and feature importance.

| Feature | Description | Importance |
| --- | --- | --- |
| Medicare/Medicaid | If the patient has active Medicare or Medicaid coverage at the time of the prediction encounter | 0.022150 |
| Has Primary Care Provider | If the patient has a Primary Care Provider indicated in Epic at the time of the prediction encounter. | 0.027619 |
| Age | The patient's age at the time of the prediction encounter. | 0.132686 |
| Gender - Male | If the patient's gender is Male at the time of the prediction encounter. | 0.036439 |
| Single | If the patient relationship status is indicated as single at the time of the prediction encounter. | 0.018067 |
| Married or significant other | If the patient relationship status is indicated as married or has a significant other at the time of the prediction encounter. | 0.020761 |
| Divorced, separated, or widowed | If the patient relationship status is indicated as divorced, separated, or widowed at the time of the prediction encounter. | 0.015483 |
| Has Coronary Artery Disease (CAD) | The patient is active on the Coronary Artery Disease (CAD) registry at the time of the prediction encounter. | 0.008645 |
| Has Congestive heart failure (CHF) | The patient is active on the Congestive heart failure (CHF) registry at the time of the prediction encounter. | 0.003730 |
| Has Chronic kidney disease (CKD) | The patient is active on the Chronic kidney disease (CKD) registry at the time of the prediction encounter. | 0.005192 |
| Has Chronic liver disease (CLD) | The patient is active on the Chronic liver disease (CLD) registry at the time of the prediction encounter. | 0.004033 |
| Has Chronic obstructive pulmonary disease (COPD) | The patient is active on the Chronic obstructive pulmonary disease (COPD) registry at the time of the prediction encounter. | 0.013216 |
| Has diabetes | The patient is active on the diabetes registry at the time of the prediction encounter. | 0.013223 |
| Has Human Immunodeficiency Virus (HIV) | The patient is active on the Human Immunodeficiency Virus (HIV) registry at the time of the prediction encounter. | 0.000007 |
| Has obesity | The patient is active on the obesity registry at the time of the prediction encounter. | 0.024823 |
| Chronic opioids | The patient is active on the chronic opioids registry at the time of the prediction encounter. | 0.003502 |
| Asian | The patient primary race is indicated as Asian | 0.000376 |
| Black or African American | The patient primary race is indicated as Black or African American | 0.010433 |
| White or Caucasian | The patient primary race is indicated as White or Caucasian | 0.017762 |
| Hispanic or Latino | The patient primary race is indicated as Hispanic or Latino | 0.001588 |
| Alcohol use | If the social history in the last 3 years from time of prediction encounter indicates alcohol use. | 0.025474 |
| Alcohol ounce (oz.) per week | Maximum consumption of reported ounces per week of alcohol in the last 3 years from time of prediction encounter. | 0.024881 |
| Tobacco use | If the social history in the last 3 years from time of prediction encounter indicates tobacco use. | 0.026711 |
| Illicit drug use | If the social history indicates use of illicit drugs before the time of prediction encounter. | 0.007396 |
| Access to firearms | If the patient indicated that they have access to firearms on the Behavioral Health Access form in the last 1 year from time of prediction encounter. | 0.005630 |
| Hopelessness/Helplessness | If the patient indicated Hopelessness/Helplessness on the Behavioral Health Access form in the last 1 year from time of prediction encounter. | 0.013006 |
| Attempt or self-injury composite | If the patient has indicated that they have a current/recent/history of suicide attempt or self-injury on the Behavioral Health Access form in the last 1 year from time of prediction encounter. | 0.006491 |
| Patient Health Questionnaire-9 (PHQ9) total score | The total score for the most recent Patient Health Questionnaire-9 (PHQ9) assessment which is not more than 1 year from the time of prediction encounter. | 0.038088 |
| Generalized Anxiety Disorder (GAD) Assessment total score | The total score for the most recent Generalized Anxiety Disorder (GAD) assessment which is not more than 1 year from the time of prediction encounter. | 0.010135 |
| On antidepressants | If the patient has a medication order for a drug in the ACO ERX General Depression Medications grouper 6 months prior to the prediction encounter. | 0.025472 |
| On antipsychotics | If the patient has a medication order for a drug in the Antipsychotics pharmacy class 6 months prior to the prediction encounter. | 0.018418 |
| Behavioral health discharge in the past 72 hours | If the patient was discharged from a behavioral health unit at any Novant Health facility within 72 hours prior to the prediction encounter. | 0.001677 |
| Behavioral health discharge in the past 30 days | If the patient was discharged from a behavioral health unit at any Novant Health facility within 30 days prior to the prediction encounter. | 0.004579 |
| Behavioral health discharge in the past year | If the patient was discharged from a behavioral health unit at any Novant Health facility within the year prior to the prediction encounter. | 0.020410 |
| Major Depressive Disorder (MDD) | If the patient has had an encounter diagnosis for Major Depressive Disorder (MDD) at any time prior to the prediction encounter. | 0.019489 |
| Bipolar disorder | If the patient has had an encounter diagnosis for Bipolar disorder at any time prior to the prediction encounter. | 0.009437 |
| Schizophrenia | The patient has had an encounter diagnosis for Schizophrenia at any time prior to the prediction encounter. | 0.001398 |
| Schizoaffective | If the patient has had an encounter diagnosis for Schizoaffective at any time prior to the prediction encounter. | 0.000741 |
| Insomnia | If the patient has had an encounter diagnosis for Insomnia at any time prior to the prediction encounter. | 0.022332 |
| Personality | If the patient has had an encounter diagnosis for Personality disorder at any time prior to the prediction encounter. | 0.001843 |
| Anxiety | If the patient has had an encounter diagnosis for Anxiety at any time prior to the prediction encounter. | 0.029484 |
| Post-traumatic stress disorder (PTSD) | If the patient has had an encounter diagnosis for Post-traumatic stress disorder (PTSD) at any time prior to the prediction encounter. | 0.002371 |
| Major depressive disorder diagnosed in last 6 months | If the patient has had an encounter diagnosis for Major depressive disorder for the first time within 6 months prior to the prediction encounter. | 0.012966 |
| Bipolar disorder diagnosed in last 6 months | If the patient has had an encounter diagnosis for Bipolar disorder for the first time within 6 months prior to the prediction encounter. | 0.000796 |
| Schizophrenia diagnosed in last 6 months | If the patient has had an encounter diagnosis for Schizophrenia for the first time within 6 months prior to the prediction encounter. | 0.001182 |
| Schizoaffective diagnosed in last 6 months | If the patient has had an encounter diagnosis for Schizoaffective for the first time within 6 months prior to the prediction encounter. | 0.000071 |
| Insomnia diagnosed in last 6 months | If the patient has had an encounter diagnosis for Insomnia for the first time within 6 months prior to the prediction encounter. | 0.010209 |
| Personality diagnosed in last 6 months | If the patient has had an encounter diagnosis for Personality disorder for the first time within 6 months prior to the prediction encounter. | 0.003656 |
| Anxiety diagnosed in last 6 months | If the patient has had an encounter diagnosis for Anxiety for the first time within 6 months prior to the prediction encounter. | 0.017339 |
| Post-traumatic stress disorder (PTSD) diagnosed in last 6 months | If the patient has had an encounter diagnosis for Post-traumatic stress disorder (PTSD) for the first time within 6 months prior to the prediction encounter. | 0.000406 |
| Drug or Alcohol abuse | If the patient has had an encounter diagnosis for Drug or Alcohol abuse at any time prior to the prediction encounter. | 0.024698 |
| Drug or Alcohol abuse diagnosed in last 6 months | If the patient has had an encounter diagnosis for Drug or Alcohol abuse for the first time within 6 months prior to the prediction encounter. | 0.005854 |
| Prior suicide attempt or self-harm in last 6 months | If the patient has had an encounter diagnosis for Suicide and Intentional Self-Inflicted Injury within 6 months prior to the prediction encounter. | 0.072841 |
| Prior suicide attempt or self-harm more than 6 months ago | If the patient has had an encounter diagnosis for Suicide and Intentional Self-Inflicted Injury more than 6 months ago from the prediction encounter. | 0.014476 |
| On Benzodiazepines | If the patient has a medication order for a drug in the Benzodiazepines grouper 6 months prior to the prediction encounter. | 0.023327 |
| On Hypnotics | If the patient has a medication order for a drug in the Hypnotics pharmacy class 6 months prior to the prediction encounter. | 0.018933 |
| Outpatient Behavioral health encounter in past 72 hours | If the patient had an outpatient behavioral health encounter from unit at any Novant Health facility with certain specialties within 72 hours prior to the prediction encounter. | 0.002036 |
| Outpatient Behavioral health encounter in past 30 days | If the patient had an outpatient behavioral health encounter from unit at any Novant Health facility with certain specialties within 30 days prior to the prediction encounter. | 0.002226 |
| Outpatient Behavioral health encounter in past year | If the patient had an outpatient behavioral health encounter from unit at any Novant Health facility with certain specialties within a year prior to the prediction encounter. | 0.005396 |
| Attention deficit disorder (ADD) | If the patient has had an encounter diagnosis for Attention deficit disorder (ADD) at any time prior to the prediction encounter. | 0.008516 |
| Autism spectrum disorder (ASD) | If the patient has had an encounter diagnosis for Autism spectrum disorder (ASD) at any time prior to the prediction encounter. | 0.000802 |
| Conduct disorder | If the patient has had an encounter diagnosis for Conduct disorder at any time prior to the prediction encounter. | 0.000367 |
| Dementia | If the patient has had an encounter diagnosis for dementia at any time prior to the prediction encounter. | 0.005813 |
| Eating disorder | If the patient has had an encounter diagnosis for Eating disorder at any time prior to the prediction encounter. | 0.004055 |
| Traumatic Brain Injury (TBI) | If the patient has had an encounter diagnosis for Traumatic Brain Injury (TBI) at any time prior to the prediction encounter. | 0.004714 |
| Attention deficit disorder (ADD) diagnosed in last 6 months | If the patient has had an encounter diagnosis for Attention deficit disorder (ADD) for the first time within 6 months prior to the prediction encounter. | 0.000451 |
| Dementia diagnosed in last 6 months | If the patient has had an encounter diagnosis for Dementia for the first time within 6 months prior to the prediction encounter. | 0.002033 |
| Traumatic Brain Injury (TBI) diagnosed in last 6 months | If the patient has had an encounter diagnosis for Traumatic Brain Injury (TBI) for the first time within 6 months prior to the prediction encounter. | 0.005943 |
| Charlson Comorbidity Index (CCI) Score | The patient's Charlson Comorbidity Index (CCI) score is calculated prior to the prediction encounter. | 0.055663 |

The Receiver Operating Characteristic curve is shown in **Fig 1**. As previously mentioned, as we are more interested in evaluating the model in terms of predicting true positives rather than true negatives, we focused mainly on the area under precision recall curve (AUPRC) which avoids the influence of true negatives. The Precision-Recall curve is shown in **Fig 2**.

**Fig 1.**
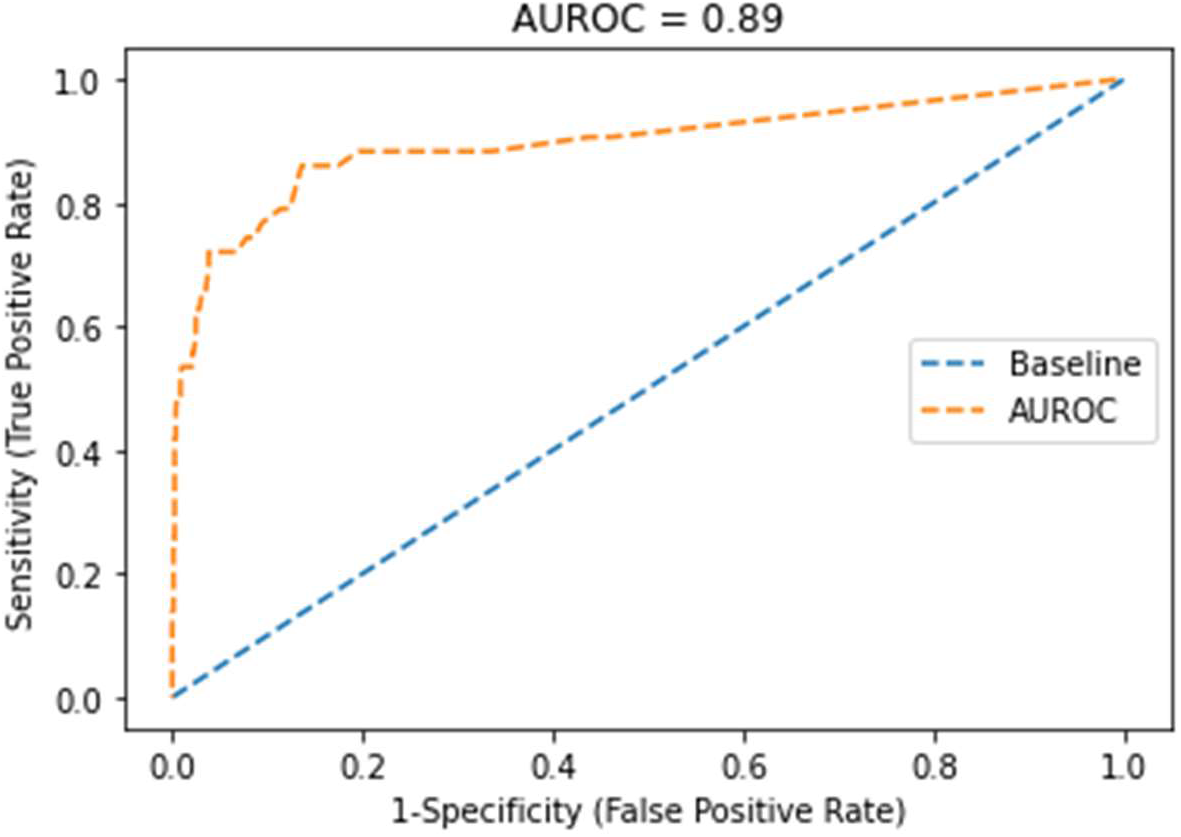
Receiver Operating Characteristic curve of the random forest model.

**Fig 2.**
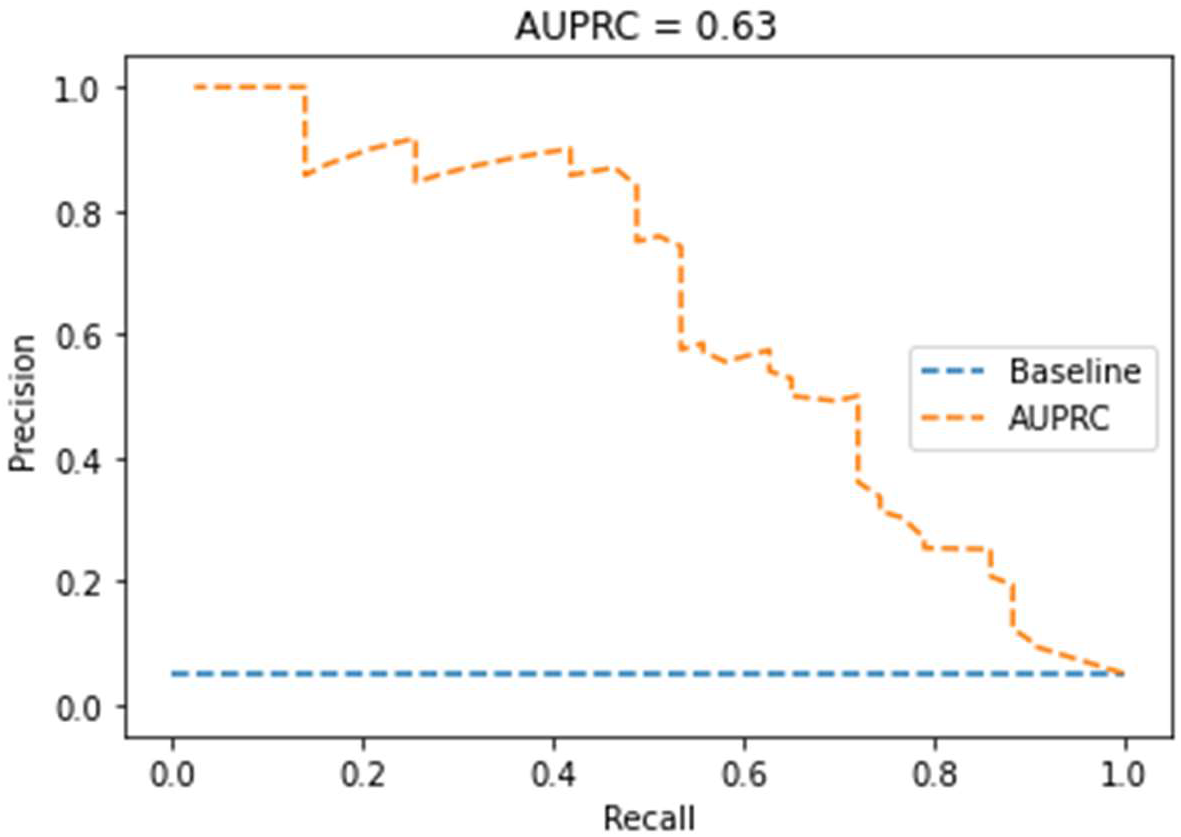
Precision-Recall curve of the random forest model.

A threshold selection process was implemented using the F1 score to select a threshold above which a patient can be considered as at high risk. As the F1 score is the harmonic mean of precision and recall, higher F1 scores indicate higher precision and recall, which in turn implies lower false positive and false negative rates. The F1 score is calculated at every threshold with intervals of 0.01 and plotted as shown in **Fig 3**. The threshold at which the F1 score peaks, at 44% risk, was selected as the ideal threshold for classifying a patient as high risk or not, though generally a plateau in F1 score occurs between approximately 30% and 60% risk. The F1 score and other performance metrics at thresholds of 30%, 44%, and 60% are listed in **Table 2**.

**Table 2.** Performance metrics for select thresholds.

|  | Threshold |  |  |
| --- | --- | --- | --- |
|  | 30% | 44% | 60% |
| **Sensitivity** | 0.65 | 0.53 | 0.41 |
| **Specificity** | 0.96 | 0.99 | 0.99 |
| **Precision** | 0.52 | 0.74 | 0.85 |
| **Negative Predictive Value** | 0.98 | 0.97 | 0.96 |
| **Accuracy** | 0.95 | 0.96 | 0.96 |
| **F1 Score** | 0.58 | 0.62 | 0.56 |

**Fig 3.**
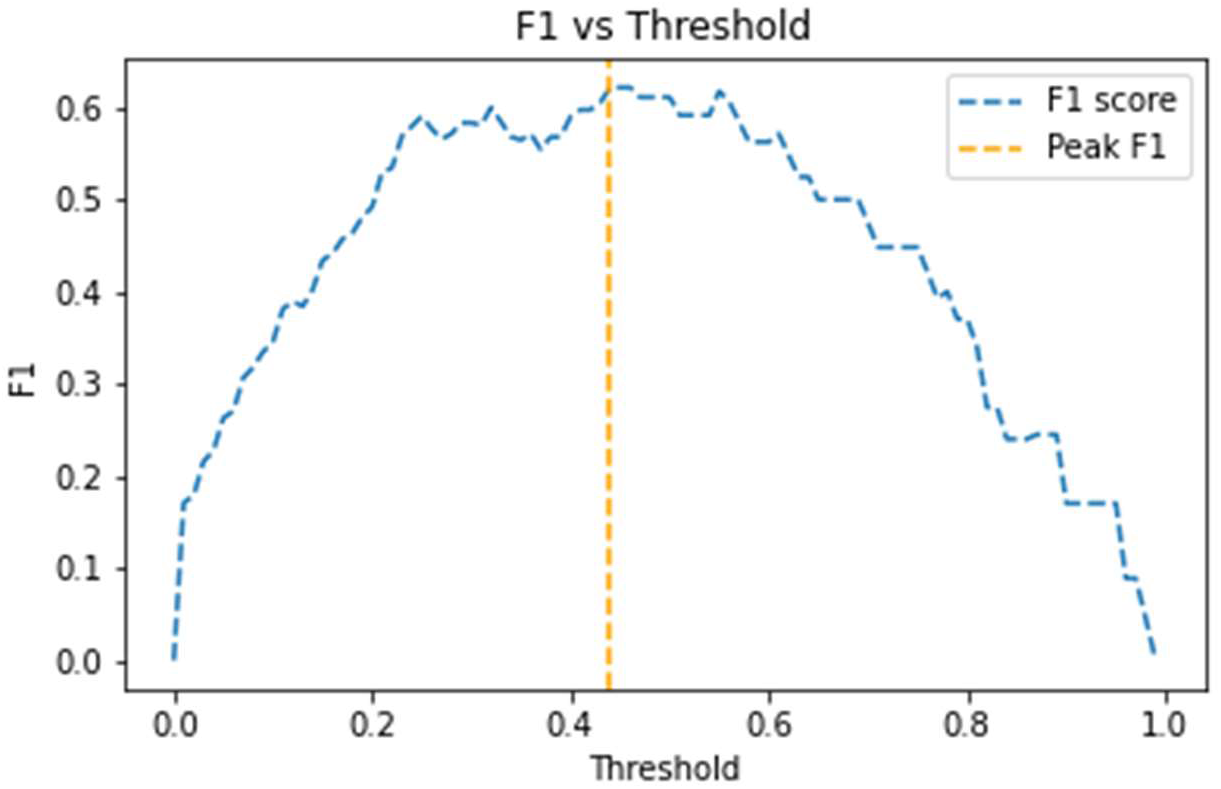
F1 score by risk threshold.

It is worth noting that, as with other probabilistic models, it is not strictly necessary to choose a single threshold for clinical intervention. The BHAR score could be used to rank patients by decreasing risk to prioritize limited resources such as behavioral health specialists to the highest risk patients first and proceed down the list in reverse rank order. Similarly, a low-medium-high categorization could be used to address the highest risk patients with the most resource intensive interventions and medium risk patients, comprising a larger proportion of patients, with less resource intensive interventions.

At all emergency departments included in this study, the C-SSRS is employed as a suicidal ideation screening tool. The C-SSRS assigns a risk level (No Risk, Low Risk, Moderate Risk, or High Risk) based on patient responses to questions about suicidal thoughts. When examining the one year period over 2021, a total of 386,130 patients were seen at a Novant Health emergency department and completed the C-SSRS screening. Of those, six individuals completed suicide within a week of the ED visit. All six individuals were assessed as No Risk based on their responses to the C-SSRS. The BHAR model would have assigned risk scores of 72%, 51%, 26%, 11%, 2%, and 0% to these six individuals. However, the C-SSRS is a very valuable tool for identifying patients ready to talk about suicidal ideation. In fact, out of the patients screened using the C-SSRS who were rated as High Risk (6,684 or 1.7%), none were identified as having completed suicide within a week of discharge, supporting that identifying patients ready to speak to someone and connecting them to those resources is a successful strategy for suicide prevention. However, an objective data-based risk score such as the BHAR score can supplement self-reported screeners by identifying patients at risk based on data in their medical record. Some patients may respond better to a more personal and gentle inquiry into whether they would like to speak to someone, whereas the C-SSRS, delivered as a standard set of questions asked to every patient, may be perceived as procedural and impersonal.

### Limitations and future work

One limitation to this work is the way in which suicide completions are labeled in the medical record and in mortality records. Oftentimes coding for accidental overdose or accidental self-inflicted injury is used when intent is not clear, and these cases would have been omitted from the training set used to train the model to identify patients at risk.

There are a few opportunities for model improvement which would be valuable enhancements to this work. One is the exploration of sexual orientation and gender identity (SOGI) data as model features. Members of the LGBTQ+ community are known to be disproportionately affected by depression and suicide (19), so inclusion of SOGI information could be reasonably expected to have value in a suicide risk model. SOGI aspects are not well documented in the Novant Health EHR at the time of this study, though efforts are underway to increase this documentation for the purposes of improving patient care and patient experience.

Natural language processing could also increase model performance by examining text communication between patients and providers, such as through MyChart, and documentation notes made by care providers during the normal course of care. One approach could be to finetune the publicly available Clinical BERT model (20) for this task.

## Data Availability

All data produced in the present study may be made available upon reasonable request to the authors after review by the Novant Health Information Governance Steering Committee.

## Acknowledgments

The authors would like to acknowledge colleagues in Novant Health Digital Products and Services who enabled this research through support of the Novant Health data warehouse and data platform. The authors extend gratitude to Sean Ganon who assisted in obtaining mortality records for the state of North Carolina and for literature review relevant to this work.

## Notes

### Competing Interest Statement

The authors have declared no competing interest.

### Funding Statement

This study did not receive any funding.

### Author Declarations

The Institutional Review Board of Novant Health, Inc gave ethical approval for this work.

### Summary of Updates

Author affiliation has been updated for Raj Patel, MD.

